# Paretic upper extremity strength at acute rehabilitation evaluation predicts motor function outcome after stroke

**DOI:** 10.1101/2021.10.05.21264572

**Authors:** Mary Alice Saltão da Silva, Christine Cook, Cathy M Stinear, Steven L Wolf, Michael R Borich

**Affiliations:** Division of Physical Therapy, Department of Rehabilitation Medicine, Emory University, Atlanta, Georgia, USA; School of Biological Sciences, Georgia Institute of Technology, Atlanta, Georgia, USA; Department of Medicine, University of Auckland, Auckland, New Zealand; Centre for Brain Research, University of Auckland, Auckland, New Zealand; Atlanta Veterans Affairs Health Care System, Center for Visual and Neurocognitive Rehabilitation, Decatur, Georgia, USA

## Abstract

**Objective:** The primary objective of this study was to retrospectively assess current care practices to determine the routinely collected measures that are most predictive of paretic upper extremity (PUE) functional outcome post-stroke in patients undergoing acute inpatient rehabilitation (AR).

**Methods:** We conducted a longitudinal chart review of patients post-stroke who received care in the Emory University Hospital system for acute hospitalization, AR, and outpatient therapy in fiscal years 2016-2018. We identified eligible patients using previously established inclusion and exclusion criteria. We extracted demographics, stroke characteristics, and longitudinal documentation of post-stroke motor function from institutional electronic medical records. Serial assessments of PUE strength were estimated using available shoulder abduction and finger extension manual muscle test documentation (E-SAFE). Estimated Action Research Arm Test (E-ARAT) was used to quantify 3-month PUE functional outcome. Metric associations were explored through correlation and cluster analyses, Kruskal-Wallis tests, classification and regression tree (CART) analysis.

**Results:** Thirty-four patients met study eligibility criteria. E-SAFE assessments performed closest to acute hospitalization day-3 (Acute E-SAFE) and upon AR admission (AR E-SAFE) were correlated with E-ARAT. Cluster analysis produced three distinct outcome groups and aligned closely to previous outcome categories. Outcome groups significantly differed in Acute E-SAFE and AR E-SAFE. Exploratory CART analysis selected AR E-SAFE to classify patient outcome with 70.6% accuracy.

**Conclusions:** Current study findings reveal that: PUE E-SAFE, measured both acutely and at AR admission, is associated with PUE motor recovery outcome; categorizations of outcome are consistent with previous studies; and predictive models can identify recovery outcome category in patients undergoing AR.

**Impact Statement:** Our findings highlight the clinical utility of SAFE as an easy-to-acquire, readily implementable screening metric. Early, intentional use of SAFE in AR settings may improve clinical decision-making, enabling therapists to deliver precision-based interventions that serve to speed or enhance recovery outcome for patients post-stroke.

## Introduction

Stroke is the leading cause of long-term adult disability worldwide.^1^ Most patients experience persistent upper extremity motor impairment.^2,3^ Recovery of paretic upper extremity (PUE) motor function is a primary determinant of functional independence in activities of daily living and quality of life.^4–8^ The majority of motor recovery occurs early after stroke, typically plateauing around 3-months post-injury, and is thought to be regulated by molecular mechanisms underlying structural and functional reorganization and the restoration of excitatory and inhibitory neurotransmitter balance within both lesioned and non-lesioned hemispheres.^2,3,9–13^ These processes together are the ingredients for spontaneous biological recovery that contribute to recovery of function in the first 3-months post-stroke (**Figure 1**).

**Figure 1.**
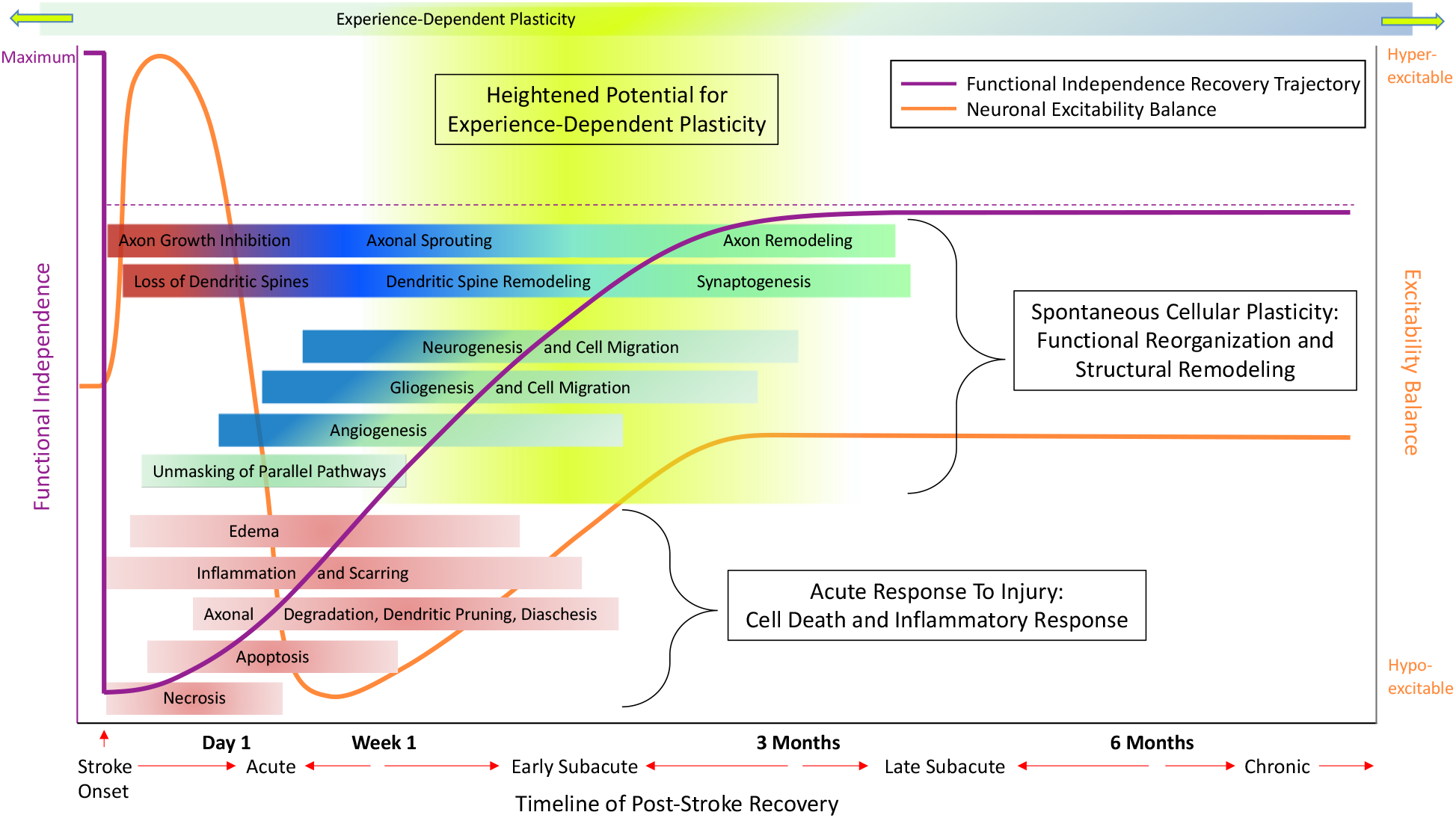
The pathophysiology of post-stroke recovery is a dynamic process. Immediately post-stroke some amount of functional independence is lost for most patients (left y-axis and purple line) due to both the extent of damage to cerebral tissues and excitability changes mediated by alterations of neurotransmitter balance (right y-axis and orange line). Acute inflammatory responses, control of cortical excitability balance (orange line), and spontaneous cellular repair mechanisms promote return of functional independence (purple line). Though experience-dependent plasticity occurs across the life span, the interaction between dynamic cellular and molecular recovery processes and neurorehabilitation offers a potentially heightened window for experience-dependent plasticity to further enhance recovery of motor function post-stroke (shaded yellow area). Adapted from Stinear and Byblow, 2014.

Therapeutic interventions that target task-specific, experience-dependent plasticity induction are the foundation of contemporary post-stroke neurorehabilitation strategies. Early therapeutic intervention promotes recovery of PUE skills such as reaching, grasping, or pinching, all of which underlie functional independence in activities of daily living.^14–16^ When these strategies are employed during a unique time of endogenous cellular repair and enhanced central nervous system reorganization, there is an interaction between cellular processes and behavioral activity.^17^ Thus, the timeframe of dynamic cellular and molecular processes offers an enticing target to augment motor recovery for patients’ post-stroke (shaded yellow area, **Figure 1**).

Critically, this period coincides with the timeframe of acute inpatient rehabilitation (AR), the setting in which the majority of post-stroke rehabilitation services and expenditures occur, thereby emphasizing the importance of delivering precision-based therapeutic interventions in AR that are effective in targeting task-specific plasticity to enhance recovery of PUE motor function.^18–21^

Early, accurate prognosis of motor outcomes would inform the delivery and specification of rehabilitative services individualized to the patient. While a codified set of measures that predict a patient’s PUE functional outcome is lacking within the US healthcare system,^22^ predictive models of stroke outcomes have been developed in healthcare systems in other countries.^23–26^ The Predict Recovery Potential (PREP2) prediction tool, developed and internally validated in New Zealand (NZ), predicts PUE motor outcomes using a combination of clinical assessments and objective neurological biomarkers.^24^ Implementation of the PREP2 prediction tool into clinical practice in NZ resulted in therapist-led modifications to clinical decision-making that were directly informed by outcome predictions.^27^ For example, therapists decreased the amount of passive movement in PUE therapy sessions for patients with good predicted outcomes.^27^ Improved therapist confidence contributed to modifications in therapeutic planning and progression, resulting in reduced lengths of inpatient hospitalization while demonstrating equivalent PUE motor outcomes at 90 days compared to when the tool was not used.^27^

NZ operates a free, public healthcare infrastructure with a unique identifier for each patient (national health index number), facilitating transfer of information with patients as they move through the continuum of post-stroke care.^28,29^ In contrast, the United States (US) healthcare system is structurally fragmented and facilities are often siloed in their patient care strategies and management systems.^30^ Therapists treating patients in AR settings in the US often lack access to therapeutic records at other time points along the care continuum. This limits knowledge of a patient’s recovery trajectory before AR but, perhaps more importantly, probably prohibits longitudinal tracking to evaluate patient outcomes. Such differences in international healthcare delivery models create a barrier to the execution of high fidelity PREP2 validation studies and may subsequently limit the generalization of findings.^18,28,31,32^ To date, two studies investigated PREP2 metrics in a healthcare system outside of NZ, but initial data collection occurred on a timeline that coincides more closely to AR admission in the US (initial strength measurements made for patients approximately 1- to 2-weeks post-stroke).^33,34^ Prediction accuracy in both studies was similar but lower than in the NZ cohort (∼60% overall vs. NZ accuracy=75%).^24,33^ The lower accuracy may be explained by only including participants admitted to inpatient rehabilitation, by omission of biomarkers of functional corticospinal tract integrity, and by delayed initial strength measurement.^33,34^ However, the lower observed accuracy also emphasizes the need to identify measures that can be collected at admission to AR that more accurately predict stroke outcome.

Accordingly, there is a need to identify measures made at later timepoints of recovery that may better predict stroke outcome in the subacute stage. There is also a need to address the inherent challenges with implementation of prediction tools reliant on early patient assessments.

Therefore, the primary objective of this study was to retrospectively assess current care practices to determine which routinely collected measures are most predictive of PUE functional outcome post-stroke in patients undergoing AR. We chose an observational, retrospective study design because we were most interested in identifying rapidly implementable, standard-of-care metrics that could predict PUE outcomes and guide care in AR settings. We hypothesized that measures routinely collected as part of standard clinical care post-stroke would predict PUE outcome category.

## Methods

### Study Population and Eligibility Criteria

We conducted a longitudinal retrospective chart review of a subset of patients admitted with a primary diagnosis of stroke who received care in the Emory University Hospital (EUH) system, a representative, urban, academic, comprehensive stroke care center in the US, between September 1, 2016 and August 31, 2018. We selected cases using previously established inclusion and exclusion criteria.^24^ Major inclusion criteria included the following: first ever or recurrent, ischemic or intracerebral hemorrhagic (ICH) stroke; new upper extremity weakness beginning at or after current stroke onset; age of ≥18 years.^24^ In addition, individuals were required to have remained within the EUH system for acute hospitalization, acute inpatient rehabilitation at Emory Rehabilitation Hospital (ERH), and Emory outpatient therapy through at least 90 days post-stroke to permit longitudinal assessment of PUE recovery outcomes and reduce the heterogeneity of post-stroke care for the study cohort across the continuum of recovery. A single patient did not begin outpatient care until approximately 1-year post-stroke (428 days) but was included in the analysis. Since the plateau of motor recovery typically occurs around 90 days post-stroke, measurement of functional outcome at a time point after that time point would enable an approximation of day-90 PUE functional outcome. Lastly, patients were required to have received diagnostic magnetic resonance imaging (MRI) while at EUH as a separate inclusion criterion associated with a parallel investigation. Patients were identified from an existing stroke database and by reviewing records of ERH admits and Emory outpatient records. This study received Emory University Institutional Review Board approval and patient consent was waived.

### Data Extraction and Analysis

Clinical metrics including demographic information, stroke characteristics, care continuum metrics, and provider documentation of post-stoke motor function were extracted from Cerner Powerchart, the institutional electronic medical record (EMR) system of the Emory Healthcare system. Data were extracted longitudinally across acute hospitalization, acute inpatient rehabilitation, and outpatient therapy through at least 90 days post-stroke.

Identified stroke characteristics extracted included: stroke type, location, imaging obtained, and stroke severity as measured by the National Institutes of Health Stroke Scale (NIHSS).^35^ If the NIHSS was measured more than once, the assessment performed closest to inpatient day-3 was used. Care continuum metrics included length of hospitalizations, duration in outpatient therapy, and time to therapy evaluation(s). Provider documentation of PUE strength and post-stroke disability included manual muscle test scores, sensation, coordination, language impairments, and measures of mobility. These metrics were recorded serially by different providers within the care continuum including physicians, physical therapists, occupational therapists, and speech language pathologists (**Table 1**).

**Table 1.** Patient demographics, comorbidities, and stroke data.

| Demographics | Median (range)/<br>Count (% total) |
| --- | --- |
| Age at stroke onset, years* | 64 (36-84) |
| Race |  |
| African American | 19 (55.9%) |
| Caucasian | 15 (44.1%) |
| Sex |  |
| Female | 14 (41.2%) |
| Male | 20 (58.8%) |
| Living situation |  |
| With family | 28 (82.4%) |
| With friend/roommate | 4 (11.8%) |
| Alone | 2 (5.9%) |

| **Comorbidities** |  |
| --- | --- |
| BMI<sup>a</sup>, kg/m2* | 29 (18.9-43.2) |
| CDC BMI<sup>a</sup> classification |  |
| Normal | 9 (26.5%) |
| Overweight | 13 (38.2%) |
| Obesity (Class 1-3) | 12 (35.3%) |
| Smoker (current or former)* | 3 (8.8%) |
| Hypertension* | 24 (70.6%) |
| Hyperlipidemia* | 13 (38.2%) |
| Previous stroke* | 16 (47.1%) |
| Residual upper extremity weakness | 3 (8.8%) |
| Type II diabetes mellitus* | 12 (35.3%) |
| Cardiac history* | 7 (20.6%) |
| Chronic kidney disease* | 5 (14.7%) |
| White matter disease, any* | 24 (70.6%) |
| Mild | 12 (35.3%) |
| Moderate | 8 (23.5%) |
| Severe/Extensive | 4 (11.8%) |
| Ischemic stroke Charlson comorbidity index (/19)* | 1 (0-5) |

| Stroke Information |  |
| --- | --- |
| Type* |  |
| Ischemic | 24 (70.6%) |
| Hemorrhagic | 8 (23.5%) |
| Ischemic with hemorrhagic transformation | 2 (5.9%) |
| Stroke hemisphere* |  |
| Right | 12 (35.3%) |
| Left | 17 (50.0%) |
| Bilateral | 5 (14.7%) |
| Stroke location* |  |
| Subcortical only | 15 (44.1%) |
| Mixed cortical and subcortical | 14 (41.2%) |
| Brainstem only | 4 (11.8%) |
| Mixed subcortical and brainstem | 1 (2.9%) |
| NIHSS*<sup>b</sup>/Stroke severity distribution |  |
| Mild (0 - 4) | 6 (17.7%) |
| Moderate (5 - 15) | 17 (50.0%) |
| Severe ($\geq 16$) | 5 (14.7%) |
| Not Tested | 6 (17.7%) |
| Pre-stroke dominant upper extremity affected* | 23 (67.7%) |
| Medical intervention* |  |
| tPA<sup>c</sup> | 3 (8.8%) |
| Mechanical thrombectomy | 2 (5.9%) |
| Hemi-craniectomy | 4 (11.8%) |
| **Impairment** |  |
| E-SAFE<sup>d</sup> score (/10)*† |  |
| Acute E-SAFE<sup>d</sup> | 5 (0-8) |
| AR<sup>e</sup> E-SAFE<sup>d</sup> | 6 (0-10) |
| Sensory impairment* |  |
| Acute | 18 (52.9%) |
| AR<sup>e</sup> | 12 (35.3%) |
| Coordination impairment* |  |
| Acute | 30 (88.2%) |
| AR<sup>e</sup> | 20 (58.8%) |
| Language impairment* |  |
| Acute | 17 (50.0%) |
| AR<sup>e</sup> | 20 (58.8%) |
| IRF Sit-to-stand (/6)* |  |
| Acute | 3 (0-4) |
| AR<sup>e</sup> | 3 (0-4) |
| IRF Ambulation (/6)* |  |
| Acute | 1 (0-4) |
| AR<sup>e</sup> | 1 (0-4) |
| **Hospital and Rehabilitation Duration** |  |
| Acute hospital LOS<sup>f</sup>, days | 6.5 (1-27) |
| AR<sup>e</sup> LOS<sup>f</sup>, days | 19.5 (6-35) |
| Outpatient therapy duration, days | 88.5 (29-314) |
| Number of outpatient occupational therapy visits | 21.5 (7-54) |

(a) Body mass index (BMI); (b) National Institutes of Health Stroke Scale (NIHSS); (c) Tissue plasminogen activator (tPA); (d) Estimated SAFE (E-SAFE); (e) Acute inpatient rehabilitation (AR); (f) Length of stay (LOS)
\*Data were available to the CART analysis
† SAFE was estimated for 251/272 (92.3%) of provider evaluations examined

Shoulder abduction (SA) and finger extension (FE) manual muscle tests were used to calculate a SAFE score (/10) for each patient.^26,36,37^ If an objective SAFE score was not available in clinical documentation, an Estimated SAFE score (E-SAFE) was calculated using available assessments of PUE strength with preference given to strength of muscles with similar spinal cord segmental innervation.^38,39^ Assessments of shoulder flexion, shoulder extension, deltoid muscle strength, or proximal strength were used as alternative tests for shoulder abduction with preference given to shoulder flexion or deltoid strength. Assessments of wrist extension, grip strength, or distal strength were used as alternative tests for finger extension with preference given to wrist extensors. If the SAFE (or E-SAFE) score was documented more than once during acute hospitalization, the assessment performed closest to inpatient day-3 was used, in accordance with previous work.^40,26,36^ In the AR setting, the SAFE (or E-SAFE) score performed closest to ERH admission was used.

Additional clinical and demographic information was extracted to evaluate the potential effects of non-stroke variables on PUE motor recovery outcomes. Demographic information included prior living situation/familial support and pre-stroke UE dominance. Clinical information included mobility status as measured by the Inpatient Rehabilitation Facility Patient Assessment Instrument (IRF-PAI), comorbidities commonly correlated with stroke prevalence, and calculation of the ischemic stroke Charlson comorbidity index (ISCCI) for all patients.^41–43^

The Action Research Arm Test (ARAT) was the primary dependent variable to quantify PUE functional outcome for each patient. The ARAT is a validated, sensitive, and reliable test, commonly used in stroke-related research to measure level of upper extremity function.^44^ Due to the retrospective nature of the study design, ARAT scores were estimated from therapy documentation at approximately 90 days post-stroke in accordance with the grading criteria for each test. Estimated ARAT (E-ARAT) scoring was conducted by two licensed, clinical neurologic therapists who were otherwise blinded to study findings. Rehabilitation provider notes were evaluated in detail to extract the following measures, where available, for each patient: clinical assessments of PUE muscle and grip strength, coordination, active and passive range of motion, observational movement analysis, therapeutic activity, exercises performed, rehabilitation goals, Nine-Hole Peg Test (9HPT) and Box and Block Test (BBT) scores as compared to matched, normative values.^45–48^ Each clinician independently reviewed the EMR and determined maximal and minimal scores for each ARAT test item, creating a score range for every patient. E-ARAT for every patient was calculated by taking each clinician’s median score and averaging the two values.

### Statistical Methodology

Descriptive analyses were performed to summarize the distribution of variables of interest for the entire cohort. Non-parametric correlation analyses (Spearman’s rho, r_S_) were performed to evaluate the relationship between clinical metrics extracted and level of PUE motor function at 3-months post-stroke (E-ARAT scores). The interrater reliability of the E-ARAT scores was assessed with an intraclass correlation coefficient (ICC), calculated using a two-way mixed effects model, considering people effects to be random and item effects to be fixed.^44,49^

A k-means cluster analysis was performed using E-ARAT scores to identify PUE outcome groups similarly to hypothesis-free classification analyses conducted in previous studies.^50^ The cluster analysis was repeated using two, three, and four clusters and a minimal clinically important difference (MCID) of 12 points on the ARAT as the minimum distance between cluster centers to identify the maximum number of meaningfully different outcome groups.^50,51^ Independent-samples means comparisons were then conducted using Kruskal-Wallis tests to identify differences in clinical metrics between outcome groups.

To explore which factor(s) may predict outcome cluster group, a classification and regression tree (CART) analysis was conducted. Gini was used to maximize homogeneity of child nodes with respect to the value of the target variable. All clinical metrics including stroke characteristics, comorbidities, SAFE scores, sensation, coordination, language impairments, and measures of mobility were available as inputs using a maximum tree depth of 1, a minimum terminal node size of 3, and automated pruning to avoid over-fitting. Positive (PPV) and negative (NPV) predictive values, sensitivity, and specificity of the resulting decision tree were also calculated.

Tests were two-tailed with significance set to p<0.05. Significance values were adjusted for multiple comparisons using Bonferroni correction with a two-tailed significance level of p=0.0017 for correlation analyses (0.05/29 comparisons) and p=0.02 for t-tests (0.05/3 comparisons). All statistical analyses were conducted using IBM^®^ Statistical Package for the Social Sciences (SPSS).

## Results

34 patients (median age (range): 64 (36-84) years, female: 14) admitted to EUH with acute stroke, discharged to inpatient rehabilitation at ERH, and continued outpatient therapy at Emory during fiscal years 2016-2018 met study eligibility criteria. Patient characteristics are summarized in **Table 1**. Interrater agreement for E-ARAT scores was high (ICC=0.846, 95% CI: 0.69–0.92, p<.0005). A true SAFE score could only be calculated for 21 of the 272 (7.7%) of provider evaluations examined and were estimated for the remaining 92.3% of assessments.

### Correlation Analyses

Spearman’s correlation analyses revealed the E-SAFE assessment performed closest to inpatient day-3 during acute hospitalization (Acute E-SAFE) was correlated with E-ARAT score (r_s_=0.59, n=34, p=0.0002) **(Figure 2a)**. The median time to Acute E-SAFE assessment was 3.0 days (range=0-12 days). The E-SAFE scores taken upon admission to AR (AR E-SAFE) were also correlated with E-ARAT score (r_s_=0.73, n=34, p<0.00005) **(Figure 2b)**. The median time to AR evaluation from stroke onset was 7.0 days (range=2-27 days). The median time to E-ARAT assessment was 90.5 days (range=69-428 days). The median time to assessment not including the single patient with only 1-year follow up data was 90.0 days (range=69-149 days).

**Figure 2.**
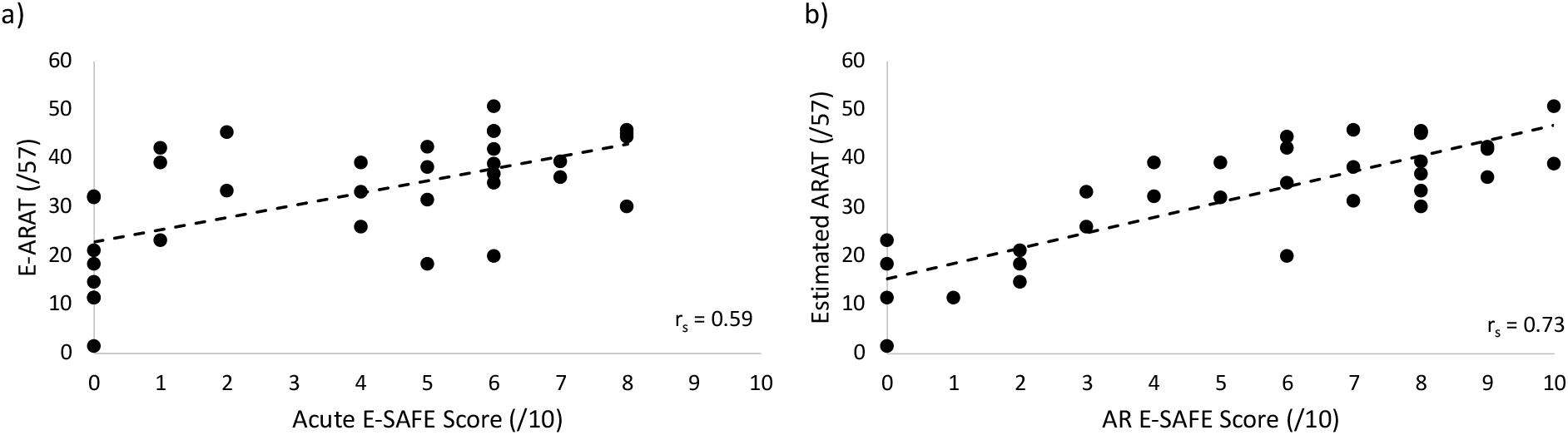
**a) Acute E-SAFE post-stroke is moderately correlated with E-ARAT score**. r_s_=0.59, n=34, ^**^p<0.01 level. **b) AR E-SAFE is strongly correlated with E-ARAT score**. r_s_=0.73, n=34, ^**^p<0.01 level.

### Cluster Analyses

Cluster analyses using two, three, and four groups all resulted in a significant difference between clusters (ANOVA p<0.001 for all cluster iterations). The three-cluster analysis produced distinct groups with centers at least 12 points (MCID) apart on the E-ARAT; however the four-cluster analysis failed to produce separation of at least one MCID between the highest scoring cluster centers (four-cluster analysis centers:1.5, 18.36, **35.39, 45.10**).^51^ Cluster cutoff scores align closely to previous predicted stroke outcome categories with PREP2^24^ (**Figure 3a)**. Based on similarities in cluster group score ranges, group nomenclature for our cohort was defined as *Good, Limited*, and *Poor*, corresponding to the ARAT score ranges identified in PREP2.^36^

**Figure 3.**
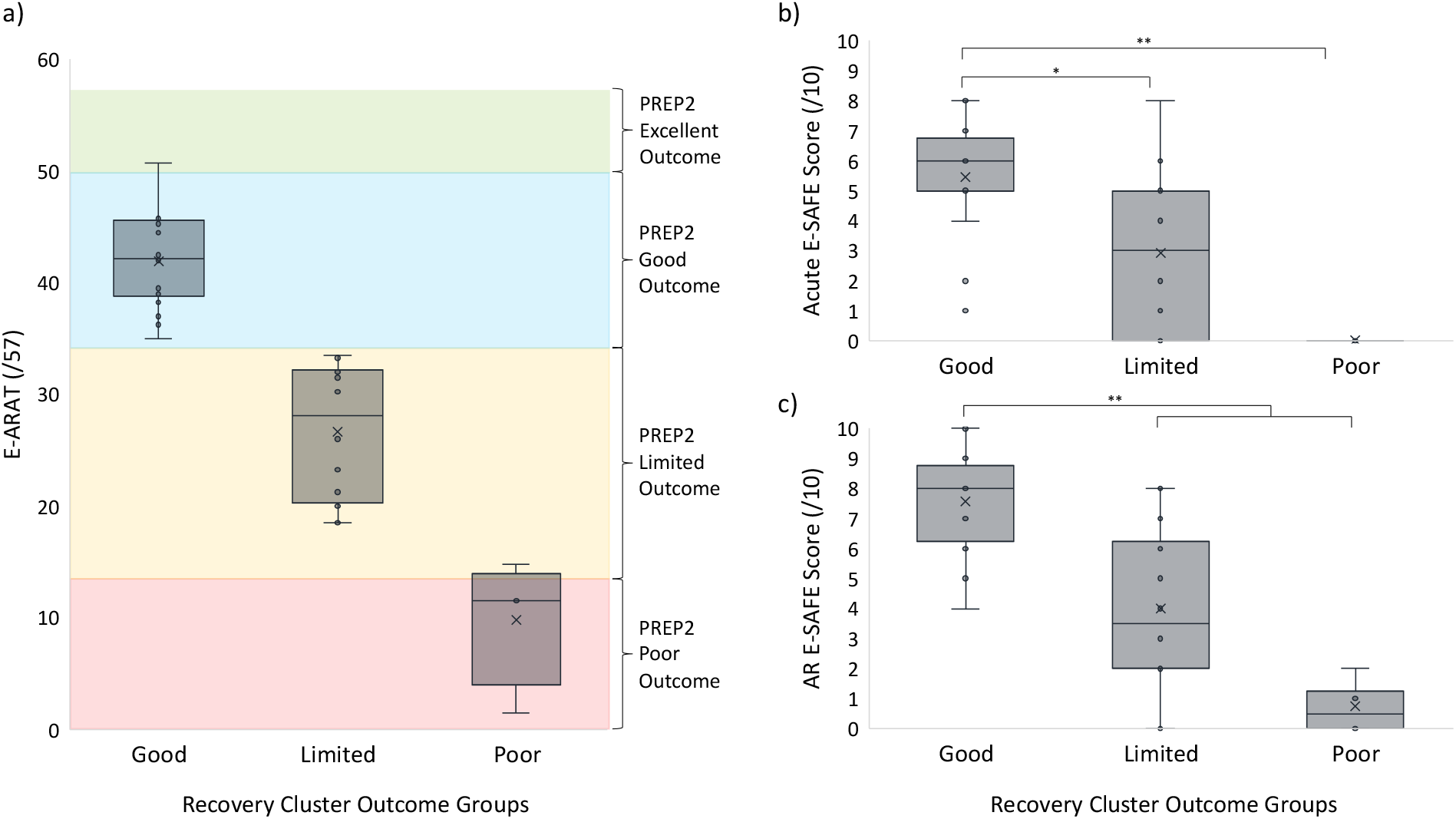
**a) Three-cluster analysis produced distinct outcome groups with centers at least 12 points (MCID) apart on the E-ARAT**. Cluster centers denoted with “x” in the figure. Three cluster cutoff scores align closely to previous predicted stroke outcome categories with PREP2, therefore group nomenclature for our cohort was defined as *Good, Limited*, and *Poor*, corresponding to previously identified ARAT score ranges.^36^ **b) Acute E-SAFE is higher for those in the *Good* outcome group over *Limited* and *Poor* outcome groups**. *Good-Limited* p=0.035, *Good-Poor* p=0.002. **c) AR E-SAFE is higher for those in the *Good* outcome group over *Limited* and *Poor* outcome groups**. *Good-Limited* p=0.007, *Good-Poor* p=0.001. All p values reported represent adjusted significance; ^*^p<0.05 level, ^**^p<0.01 level.

### Kruskal-Wallis Results

The Kruskal-Wallis test showed a significant difference in Acute E-SAFE scores between outcome groups, H(2)=14.32, p=0.001. Post-hoc pairwise comparisons revealed that Acute E-SAFE score was higher for those in the *Good* cluster than those in both the *Limited* and *Poor* clusters (*Good-Limited* median difference=3, p=0.035; *Good-Poor* median difference=6, p=0.002) (**Figure 3b, Table 2)**. The AR E-SAFE score was similarly found to be significantly different between groups, H(2)=17.47, p<0.0005. Post-hoc pairwise testing revealed that AR E-SAFE score was higher for the *Good* cluster group than both *Limited* and *Poor* groups (*Good-Limited* median difference=4.5, p=0.007; *Good-Poor* median difference=7.5, p=0.001) (**Figure 3c, Table 2)**.

**Table 2.** PUE outcome cluster group data.

| **PUE Recovery Outcome Group** | **Good** | **Limited** | **Poor** |
| --- | --- | --- | --- |
|  | Median (range) | Median (range) | Median (range) |
| Number of Patients (% total) | 18 (52.9%) | 12 (35.3%) | 4 (11.8%) |
| E-ARAT<sup>a</sup> Score (/57) | 42.25 (35-50.75) | 28.13 (18.50-33.50) | 11.50 (1.50-14.75) |
| Acute E-SAFE<sup>b</sup> Score (/10) | 6 (1-8) | 3 (0-8) | 0 (0-0) |
| AR<sup>c</sup> E-SAFE<sup>b</sup> Score (/10) | 8 (4-10) | 3.5 (0-8) | 0.5 (0-2) |
| Acute LOS<sup>d</sup> (EUH), days | 7 (2-25) | 6.5 (2-27) | 6 (1-23) |
| AR LOS<sup>d</sup> (ERH), days | 19 (6-35) | 20 (7-35) | 19.5 (17-25) |
| Outpatient therapy duration, days | 99.5 (44-314) | 82 (37-271) | 71 (29 – 157) |
| Number of outpatient visits | 20.5 (12-50) | 23.5 (11-54) | 18 (7-22) |

(a) Estimated ARAT (E-ARAT); (b) Estimated SAFE (E-SAFE); (c) Acute inpatient rehabilitation (AR); (d) Length of stay (LOS)

No clinical variables (denoted by a ^*^ in **Table 1**) differentiated the *Limited* from *Poor* outcome groups. Lengths of stay in both acute and rehabilitation hospitals were not significantly different between groups, nor was the duration of outpatient therapy or number of outpatient visits. **Table 2** contains outcome cluster patient data.

### CART Analysis

The exploratory CART analysis yielded a decision tree selecting AR E-SAFE to classify patients with 70.6% accuracy (correct classification for 24 of 34 patients) (**Figure 4)**. Patients were classified as having a *Good, Limited* or *Poor* outcome, but the decision tree failed to differentiate between *Limited* and *Poor* outcomes. For the *Good* outcome group, the PPV of the decision tree was 75.0% (18 of the 24 *Good* PUE outcome predictions were true, 6 were lower than predicted (had a *Limited* category outcome)) and the sensitivity was 100.0% (all 18 *Good* outcomes were predicted to be *Good*). The largest error was introduced for those with lower strength at admission to ERH (AR E-SAFE<4) where the accuracy was 60.0% (6 of the 10 *Limited* outcome predictions were true, 4 were lower than predicted (had a *Poor* category outcome)). For the *Limited* outcome group, the PPV was 60.0% (6 of the 10 *Limited* outcome predictions were true) and the sensitivity was 50.0% (6 of the 12 *Limited* outcomes were predicted to be *Limited*). For the *Poor* group, the PPV and sensitivity were both 0.0%. All inaccurate predictions were higher than the achieved outcome (i.e., 6 individuals predicted to be in the *Good* outcome group achieved an E-ARAT within the *Limited* outcome score range).

**Figure 4.**
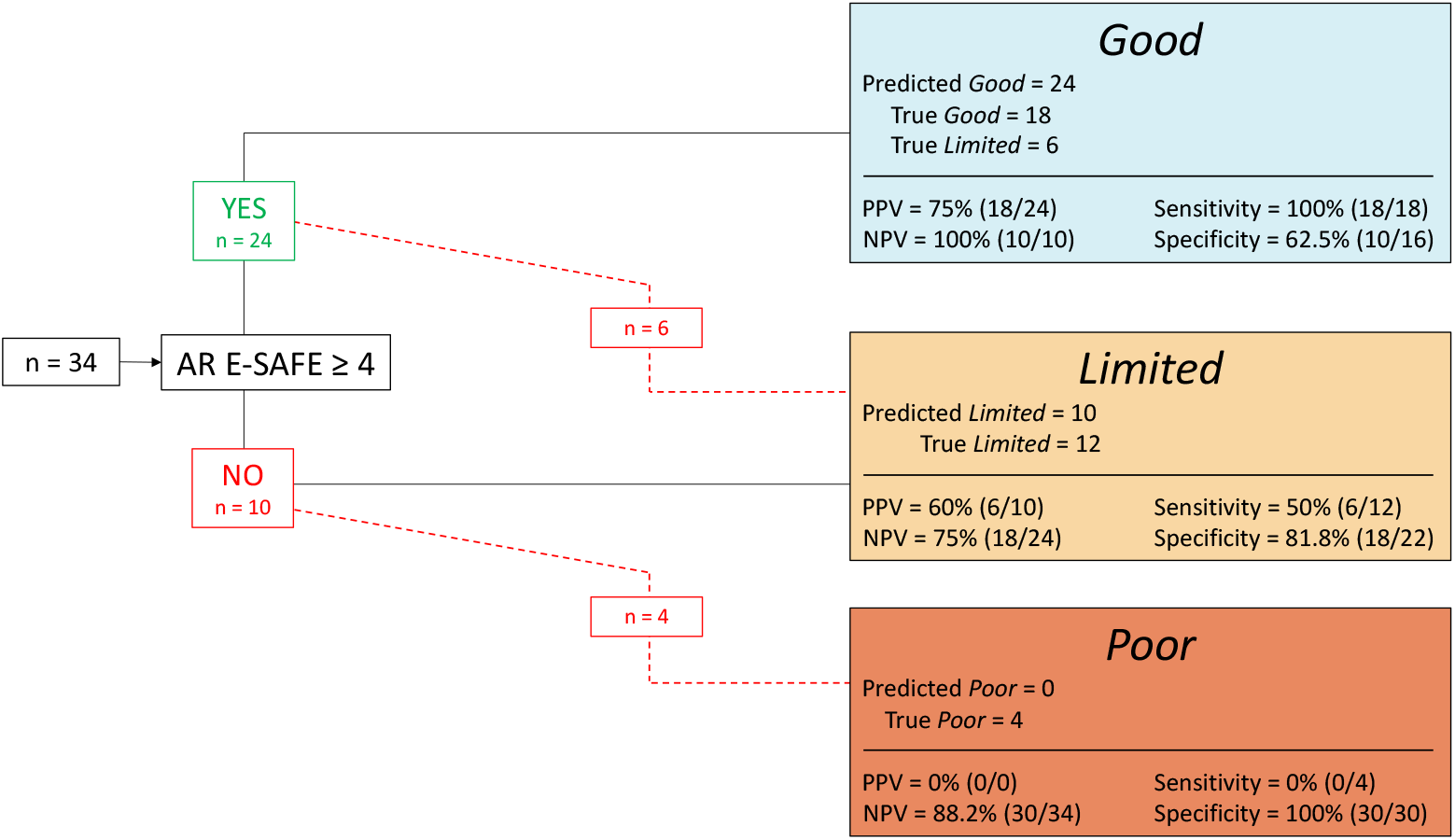
AR E-SAFE predicts PUE outcome category with 70.6% accuracy. Clinical metrics alone fail to differentiate between *Limited* and *Poor* outcomes but predict the difference between *Good* and *Limited or Poor* outcomes only. All inaccurate predictions were of a higher outcome group than achieved.

## Discussion

The current study findings reveal that PUE E-SAFE score, measured both acutely and at AR admission, are associated with PUE motor recovery outcome post-stroke, that categorization of PUE outcomes is consistent with previous studies, and that predictive models using AR E-SAFE can identify *Good* from *Limited*/*Poor* recovery outcome categories in patients undergoing AR. However, standard clinical metrics were unable to differentiate between *Limited* and *Poor* outcomes.

### Strength is strongly associated with recovery of PUE motor function

E-SAFE score emerged as the metric with the strongest association with PUE outcome at 90 days post-stroke in keeping with previous studies.^37^ This finding was true when measured early during acute hospitalization (day-3 post-stroke) and during the early subacute phase (AR admission). These findings may be unsurprising as E-SAFE score is a gross measure of baseline impairment and initial impairment has repeatedly been found to be the most powerful predictor of functional motor outcome.^52–55^ SAFE is an objective, easy-to-administer clinical metric of key muscle strength further supporting its use as a screening tool in the acute and subacute stages of recovery.^56^ In AR settings, SAFE score may be important to obtain and document as an initial screening tool for expected PUE functional recovery outcome, particularly when EMRs are not shared between care facilities or when detailed acute hospitalization records are not available.

Importantly, objective quantification is preferred to other qualitative documentation terminology commonly used by therapists (e.g., “within functional limits”) as these terms are imprecise and unclear for other providers. We observed that only 7% of provider evaluations collected objective SAFE measurements. To implement SAFE scores into routine clinical care, a structured training strategy should be considered to ensure standard measurement and documentation.^57,58^

### Categorization of PUE recovery outcome group is consistent with previous studies

Our cluster analysis resulted in E-ARAT cutoff scores which were highly consistent with categories identified with prospective ARAT assessments in PREP2.^36^ The current cohort of individuals admitted to AR had worse outcomes overall than the NZ cohort. Surprisingly, only one patient would have been classified by the PREP2 decision tool as having an *Excellent* recovery outcome (E-ARAT ≥50),^36,50^ possibly due to study selection criteria which limited our cohort to those requiring therapeutic intervention in an AR setting. Though discharge decisions are multifactorial and not solely dependent on PUE status, it may be the case that most individuals that would be predicted to have an *Excellent* PUE outcome are discharged to home from acute hospitalization rather than to AR, reflecting a robust filter within the US healthcare system which reserves the resource of AR for those who require a higher level of therapeutic intervention.

### Predictive models can identify *Good* from *Limited*/*Poor* recovery outcome categories

In our cohort, functional PUE recovery after stroke was estimated with 70% accuracy using AR E-SAFE alone. The E-SAFE score cutoff selected by the CART analysis in our decision tree is one point lower than in the PREP2 prediction tool (E-SAFE ≥4 vs. SAFE ≥5).^36^ We may expect that strength cutoff scores would decrease with time with time post-stroke as it should reflect progression of recovery. Unlike in PREP2 studies, if little to no PUE strength is available by the second week post-stroke (AR E-SAFE<4), our data suggest that a patient will achieve less than a *Good* recovery outcome without the need for further outcome potential clarification by assessing motor evoked potential status. However, a higher AR E-SAFE score does not guarantee a *Good* recovery outcome (PPV=75%). These preliminary data show promise for the creation and validation of a predictive tool with clinical utility at admission to AR in the US healthcare system but further research will be necessary to ensure these findings remain valid within a larger patient cohort.

Two studies have evaluated PREP2 metrics in healthcare settings outside of NZ and gathered initial SAFE scores in a timeline consistent with that of AR E-SAFE measurement in our cohort.^33,34^ Interestingly, both studies reported initial SAFE score medians for Limited and Poor outcome groups of <4, suggesting that similar modification of cutoff scores may be necessary to achieve optimal accuracy in new health care settings and patient populations.^33,34,59,60^ Because SAFE score continues to demonstrate strong predictive potential and has been successfully implemented in both New Zealand and Sweden, future studies should seek first to validate existing prediction tools, with high fidelity to the original metrics and timeline, with the knowledge that cutoff score modification may be necessary.^57–61^

The absence of any clinical variable that enabled statistical differentiation of *Limited* and *Poor* outcome groups suggests that future studies may need to employ distinctive metrics which quantify CST structural integrity to facilitate delineation of predictions for those with initially lower levels of PUE strength. Previous research employed high-resolution MR-based diffusion-weighted imaging biomarkers to contribute to functional outcome prediction.^24,26^ While high-resolution imaging is not standard of care in the US, lower resolution clinical neuroimaging is routinely used to diagnose stroke. Future studies should seek to leverage diagnostic imaging to identify biomarkers of CST structural integrity as they may provide a feasible option to assist in distinguishing *Limited* and *Poor* PUE functional motor outcomes.

### Limitations

This retrospective study design has strengths and limitations. The most significant limitation of the retrospective design is that it necessitated estimation of most SAFE scores and estimation of ARAT performance for all patients. This approximation likely introduces some measurement error to current findings though E-ARAT scores demonstrated excellent inter-rater reliability between experienced neurologic therapists. Further, our retrospective data provided access to current standards of clinical care and recovery outcomes within the study cohort, thus yielding a dataset that may be a more accurate representation of the recovery experience of individuals post-stroke. An additional limitation to the retrospective design is that we were unable to control for differences in the content of therapy provided at each stage of post-stroke care. However, individuals who receive therapy in the same rehabilitation settings should receive a similar dosage and type of therapeutic intervention, thereby reducing heterogeneity. Further, patients in our cohort had statistically similar therapy duration across the continuum of care. Lastly, the generalizability of current findings may also be limited due to the small sample size and an uneven distribution of individuals across categories that resulted in underrepresentation of patients in both *Limited* and *Poor* outcome categories. Though our exploratory CART analysis yielded a decision tree that accurately predicts outcome for 70% of individuals, the small sample size and category distribution may lead to overfitting of the model. Though results are promising, further investigation and validation using a larger sample size will be necessary.

## Conclusions

Tailoring therapeutic intervention to expected motor outcomes is a common theme in neurorehabilitation, yet there remains an opportunity to increase the personalization of treatment and enable recovery to an individual’s physiologic potential. Our findings suggest that patients who undergo AR post-stroke demonstrate heterogeneous levels of impairment and functional outcomes while highlighting the clinical utility of the SAFE score as a simple, easy-to-acquire, readily implementable screening metric that could guide clinical decision-making in AR. Exploratory predictive modeling suggests PUE functional outcome after stroke may be accurately predicted using AR SAFE score. In an era of precision medicine, the early and intentional use of these clinically-feasible metrics may allow for improved care plan development and optimized allocation of rehabilitation resources. Taken together, these observations support the notion that clinical information routinely collected after stroke is associated with level of recovery of PUE function and, if optimized, have the potential to inform and improve the delivery of therapeutic interventions post-stroke.

## Data Availability

The data that support the findings of this study are available upon reasonable request from the corresponding author, MB.

## Acknowledgments

The authors would like to acknowledge Amulya Noone for assistance with data extraction and organization and Theresa McLaughlin, MOT, OTR-L, CBIS for assistance with ARAT scoring.

